# Diligent medical activities of a publicly designated medical institution for infectious diseases pave the way for overcoming COVID-19: A positive message to people working at the cutting edge

**DOI:** 10.1101/2020.05.19.20107490

**Authors:** Tatsuya Nagano, Jun Arii, Mitsuhiro Nishimura, Naofumi Yoshida, Keiji Iida, Yoshihiro Nishimura, Yasuko Mori

**Affiliations:** Division of Respiratory Medicine, Department of Internal Medicine, Kobe University Graduate School of Medicine, 7-5-1 Kusunoki-cho, Chuo-ku, Kobe, Hyogo 650-0017, Japan.; Division of Clinical Virology, Center for Infectious Diseases, Kobe University Graduate School of Medicine, 7-5-1 Kusunoki-cho, Chuo-ku, Kobe, Hyogo 650-0017, Japan.; Division of Cardiovascular Medicine, Department of Internal Medicine, Kobe University Graduate School of Medicine, 7-5-1 Kusunoki-cho, Chuo-ku, Kobe, Hyogo 650-0017, Japan.; Division of Diabetes and Endocrinology, Hyogo Prefectural Kakogawa Medical Center, 203 Kanno, Kanno-cho, Kakogawa, Hyogo 675-8555, Japan.

## Abstract

The analysis of systematically collected data for COVID-19 infectivity and death rates has revealed in many countries around the world a typical oscillatory pattern with a 7-days (circaseptan) period. Additionally, in some countries the 3.5-days (hemicircaseptan) and 14-days periodicities have been also observed. Interestingly, the 7-days infectivity and death rates oscillations are almost in phase, showing local maxima on Thursdays/Fridays and local minima on Sundays/Mondays. These observations are in stark contrast with a known pattern, correlating the death rate with the reduced medical staff in hospitals on the weekends. One possible hypothesis addressing these observations is that they reflect a gradually increasing stress with the progressing week, which can trigger the maximal death rates observed on Thursdays/Fridays. Moreover, assuming the weekends provide the likely time for new infections, the maximum number of new cases might fall again on Thursdays/Fridays. These observations deserve further study to provide better understanding of the COVID-19 dynamics.

---

Severe acute respiratory syndrome coronavirus 2 (SARS-CoV-2), which causes coronavirus disease 2019 (COVID-19), is highly infectious and has spread worldwide. An important factor compounding spread is the infection of medical staff with SARS-CoV-2, which threatens the collapse of the very institutions required to treat COVID-19. The possibility of virus transmission from patients with COVID-19 to medical staff is thus of primary concern. Asymptomatic COVID-19 carriage among hospital staff could also conceivably act as a potent source of ongoing transmission^1^. Here we show that, surprisingly, none of the medical staff working at a hospital with COVID-19 patients had IgG antibodies for SARS-CoV-2, indicating that transmission from patients to medical staff did not occur in these medical workers. These results show that standard preventive measures against infectious diseases can prevent SARS-CoV-2 exposure in medical staff, and should greatly encourage medical practitioners at the front line of this pandemic.

## Methods

On May 1, 7 and 8, 2020, sera were collected from medical staff at the Hyogo Prefectural Kakogawa Medical Center, which has 353 beds and is one of 55 publicly designated medical institutions for infectious diseases—including Ebola, smallpox, plague, tuberculosis, severe acute respiratory syndrome (SARS) and Middle East respiratory syndrome (MERS)—in Japan. The institutional review boards of Kobe University Hospital and Hyogo Prefectural Kakogawa Medical Center approved the study, and written informed consent was obtained from all participants. IgG antibodies for SARS-CoV-2 in each serum sample were analyzed by immunochromatographic test (INNOVITA, Hebei, China).

## Results

Hyogo Prefectural Kakogawa Medical Center began accepting patients with COVID-19 on March 11, 2020 (Fig.1). On May 1, 7 and 8, 2020, 39 (7 in ICU), 21 (8 in ICU) and 19 (7 in ICU) patients with COVID-19 were hospitalized, respectively. A total of 509 medical staff were assigned to work on these cases. The 509 medical staff consisted of 88 men and 421 women with a median age of 39 (18 to 66) years. They were 77 doctors, 310 nurses, 1 pharmacist, 20 radiology technicians, 19 laboratory medical technologists and 82 medical assistants. 115, 18, and 72 worked in the ICU, the ambulatory unit for patients with fever and the ward for patients with COVID-19, respectively. The median time from contact with patients with COVID-19 to sera collection was 23 days. All medical staff wore masks in the hospital and performed hand disinfection with alcohol after each procedure. When they collected samples for PCR or came into contact with PCR-positive patients, they wore full personal protective equipment. All sera from the medical staff were negative for IgG antibodies, indicating that none of the staff had been infected with SARS-CoV-2 despite their close contact with the patients. Sera from patients 14 days or more after onset of COVID-19 were used as a positive control and showed a 100% (10/10) positive rate for IgG.

**Figure 1.**
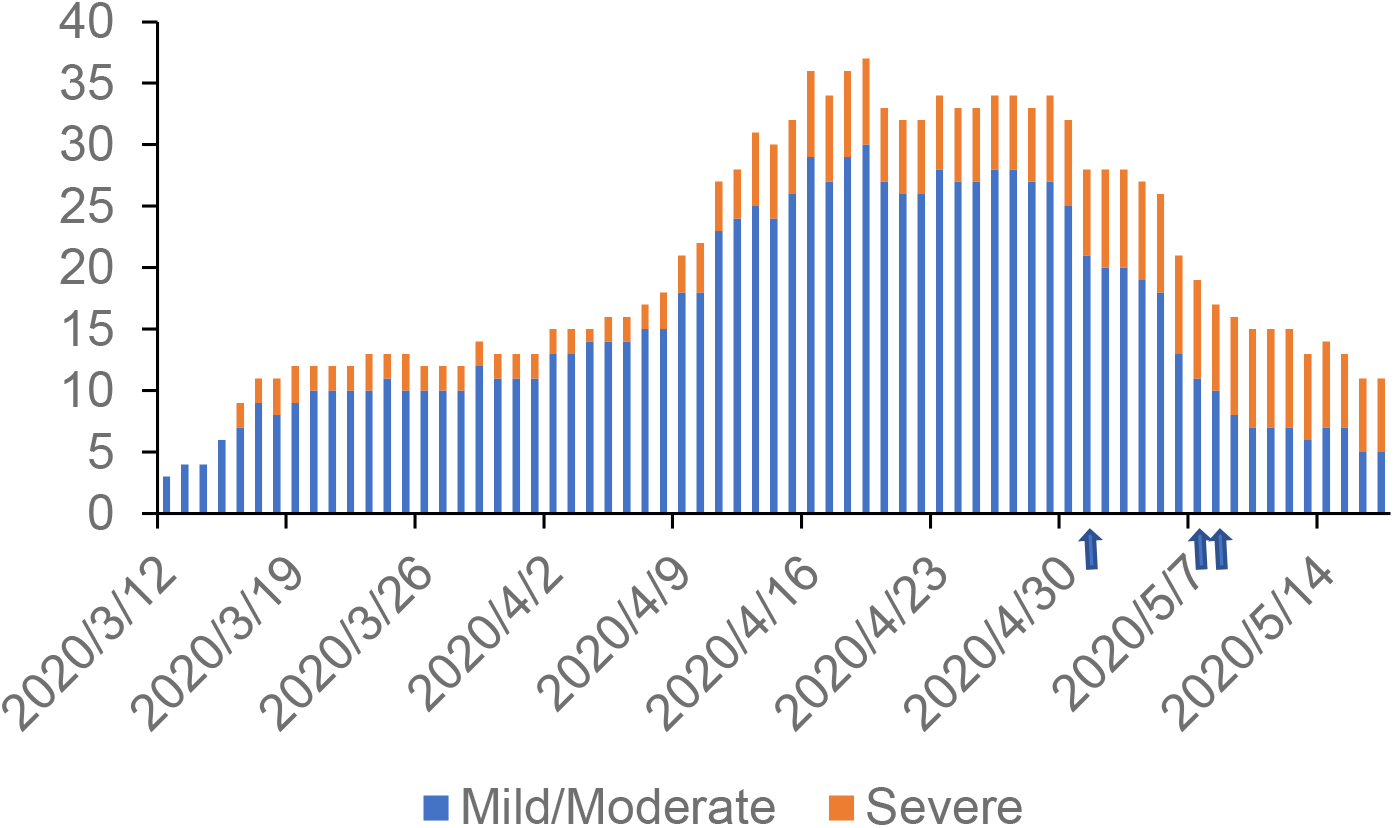
Number of hospitalized patients with COVID-19 in indicated period. Arrow, the date of collection of sera.

## Discussion

Despite the hard work of brave medical workers around the world, many patients continue to die of COVID-19^2^. Unfortunately, not a few medical staff members themselves have died from nosocomial infections of SARS-CoV-2. The medical staff at the Kakogawa Medical Center have been in contact with COVID-19 patients for up to 53 days, but so far there have been no incidents suspicious for nosocomial infection. Our surprising results suggest that standard preventive measures, if strictly followed, can prevent SARS-CoV-2 exposure in medical practitioners. For the first time, this study provides supporting evidence suggesting the usefulness of standard precautions against SARS-CoV-2. Although all medical workers naturally fear SARS-CoV-2 exposure, we believe the current results could help alleviate their anxiety, and could provide courage and inspiration for their fight against COVID-19.

## Data Availability

The availability of all data referred to in the manuscript.

## Acknowledgement

We thank Kazuro Sugimura MD, PhD (Executive Vice President, Kobe University) for his full support to promote this study. Yukiya Kurahashi, Lidya Handayani Tjan, Zhenxiao Ren, Anna Lystia Poetranto, Salma Aktar, Jing Rin Huang, and Silvia Sutandhio (Division of Clinical Virology, Center for Infectious Diseases, Kobe University Graduate School of Medicine) supported the immunochromatographic test. We express our sincere gratitude for cooperation and participation of staffs of Hyogo Prefectural Kakogawa Medical Center.

## Author Contributions

Drs. Nagano and Mori had full access to all of the data in the study and take responsibility for the integrity of the data and the accuracy of the data analysis. Drs Nagano, Arii, M. Nishimura and equally contributed to this work.

Concept and design: Nagano, Y. Nishimura, Mori.

Acquisition, analysis, or interpretation of data: Nagano, Arii, M. Nishimura, Yoshida, Iida, Y. Nishimura, Mori.

Drafting of the manuscript: Nagano, Arii, M. Nishimura, Mori.

Critical revision of the manuscript for important intellectual content: Nagano, Arii, M. Nishimura, Yoshida, Iida, Y. Nishimura, Mori.

Statistical analysis: Nagano, Arii, M. Nishimura,.

Administrative, technical, or material support: Arii, M. Nishimura

Supervision: Iida, Y. Nishimura, Mori.

## Reference

1. Black JRM, Bailey C, Przewrocka J, Dijkstra KK, Swanton C. COVID-19: the case for healthcare worker screening to prevent hospital transmission. Lancet. 05 2020;395(10234):1418–1420. doi:10.1016/S0140-6736(20)30917-X

2. Sanders JM, Monogue ML, Jodlowski TZ, Cutrell JB. Pharmacologic Treatments for Coronavirus Disease 2019 (COVID-19): A Review. JAMA. Apr 2020;doi:10.1001/jama.2020.6019

